# Adenomas from individuals with pathogenic biallelic variants in the *MUTYH* and *NTHL1* genes demonstrate base excision repair tumour mutational signature profiles similar to colorectal cancers, expanding potential diagnostic and variant classification applications

**DOI:** 10.1101/2024.08.08.24311713

**Authors:** Romy Walker, Jihoon E. Joo, Khalid Mahmood, Mark Clendenning, Julia Como, Susan G. Preston, Sharelle Joseland, Bernard J. Pope, Ana B. D. Medeiros, Brenely V. Murillo, Nicholas Pachter, Kevin Sweet, Allan D. Spigelman, Alexandra Groves, Margaret Gleeson, Krzysztof Bernatowicz, Nicola Poplawski, Lesley Andrews, Emma Healey, Steven Gallinger, Robert C. Grant, Aung K. Win, John L. Hopper, Mark A. Jenkins, Giovana T. Torrezan, Christophe Rosty, Finlay A. Macrae, Ingrid M. Winship, Daniel D. Buchanan, Peter Georgeson

## Abstract

**Background:** Colorectal cancers (CRCs) from people with biallelic germline likely pathogenic/pathogenic variants in *MUTYH* or *NTHL1* exhibit specific single base substitution (SBS) mutational signatures, namely combined SBS18 and SBS36 (SBS18+SBS36), and SBS30, respectively. The aim was to determine if adenomas from biallelic cases demonstrated these mutational signatures at diagnostic levels.

**Methods:** Whole-exome sequencing of FFPE tissue and matched blood-derived DNA was performed on 9 adenomas and 15 CRCs from 13 biallelic *MUTYH* cases, on 7 adenomas and 2 CRCs from 5 biallelic *NTHL1* cases and on 27 adenomas and 26 CRCs from 46 non-hereditary (sporadic) participants. All samples were assessed for COSMIC v3.2 SBS mutational signatures.

**Results:** In biallelic *MUTYH* cases, SBS18+SBS36 signature proportions in adenomas (mean±standard deviation, 65.6%±29.6%) were not significantly different to those observed in CRCs (76.2%±20.5%, *p-value*=0.37), but were significantly higher compared with non-hereditary adenomas (7.6%±7.0%, *p-value*=3.4x10^-4^). Similarly, in biallelic *NTHL1* cases, SBS30 signature proportions in adenomas (74.5%±9.4%) were similar to those in CRCs (78.8%±2.4%) but significantly higher compared with non-hereditary adenomas (2.8%±3.6%, *p-value*=5.1x10^-7^). Additionally, a compound heterozygote with the c.1187G>A p.(Gly396Asp) pathogenic variant and the c.533G>C p.(Gly178Ala) variant of unknown significance (VUS) in *MUTYH* demonstrated high levels of SBS18+SBS36 in four adenomas and one CRC, providing evidence for reclassification of the VUS to pathogenic.

**Conclusions:** SBS18+SBS36 and SBS30 were enriched in adenomas at comparable proportions observed in CRCs from biallelic *MUTYH* and biallelic *NTHL1* cases, respectively. Therefore, testing adenomas may improve the identification of biallelic cases and facilitate variant classification, ultimately enabling opportunities for CRC prevention.

## INTRODUCTION

Identifying people who have an increased risk of developing colorectal cancer (CRC), including people with a hereditary CRC or polyposis syndrome, provides important opportunities for cancer prevention. Individuals with homozygous or compound heterozygous likely pathogenic or pathogenic (LP/P) variants in the base excision repair genes *MUTYH* [1] and *NTHL1* [2] (i.e., biallelic cases) predispose to the development of multiple pre-cancerous adenomas in the colon (adenomatous polyposis), CRC and a spectrum of extra-colonic cancers [2,3].

The application of tumour mutational signature profiling to identify hereditary cancer syndromes related to DNA repair defects has been highlighted [4]. Single base substitution (SBS) and insertion/deletion mutational signatures in CRC have been shown to be accurate predictors of Lynch syndrome and biallelic germline LP/P variants in *MUTYH* [5]. In particular, the combination of SBS18 and SBS36 (SBS18+SBS36) can accurately identify those with germline biallelic *MUTYH* LP/P variants [5,6], while for *NTHL1*, the SBS30 mutational signature has been identified in CRCs from those with biallelic *NTHL1* LP/P variants [7]. Moreover, we previously identified two recurrent somatic mutations, namely the *KRAS* c.34G>T p.(Gly12Cys) and the *PIK3CA* c.1636C>A p.(Gln546Lys) mutations, that were strongly enriched in CRCs from biallelic *MUTYH* cases compared with CRCs from non-hereditary/sporadic cases (*KRAS*: *p-value*=1.4x10^-6^, *PIK3CA*: *p-value*=3.4x10^-4^) [6].

A further application of tumour mutational signature profiling is to aid variant classification. Previously, we have shown the presence of elevated levels of SBS18+SBS36 in CRCs provided evidence for an LP/P classification for the germline *MUTYH* variants c.1141G>T p.(Gly381Trp) and c.577-5A>G, where the second allele of *MUTYH* harboured an LP/P variant [6]. Alternatively, the absence of high levels of SBS18+SBS36 in CRCs supported a benign classification for *MUTYH* variants c.912C>G p.(Ser304Arg), c.821G>A p.(Arg274Gln), c.925C>T p.(Arg309Cys) and c.1431G>C p.(Thr477Thr) [6].

While these genomic features have been shown to be effective with CRC-derived data, there are important implications that could be facilitated by the ability to utilise mutational signature profiling in pre-cancerous adenomas namely: 1) identifying biallelic cases early before they develop cancer, including guiding surgical versus endoscopic management decision making, 2) enable pre-emptive genetic counselling and guide patient management strategies through risk assessment, 3) indicate if a second “unidentified” LP/P variant is present in monoallelic LP/P variant carriers, and 4) provide evidence for pathogenicity for variants of uncertain significance (VUS). The aim of this study was to profile and compare the SBS18+SBS36 and SBS30 mutational signatures in adenomas and CRCs from biallelic *MUTYH* and biallelic *NTHL1* cases, respectively, with sporadic adenomas and CRCs from participants without a hereditary CRC/polyposis syndrome to determine their discriminatory potential and ability to inform variant classification.

## MATERIAL AND METHODS

### Study cohort

Participants were men and women recruited to one of the following studies: 1) Applying Novel Genomic approaches to Early-onset and suspected Lynch Syndrome colorectal and endometrial cancers (ANGELS, n=4), 2) Colorectal Cancer Family Registry (CCFR, n=21) or 3) Genetics of Colonic Polyposis Study (GCPS, n=5) who were identified to have either germline biallelic *MUTYH* or germline biallelic *NTHL1* LP/P variants from clinical diagnostic or research genetic testing. Formalin-fixed paraffin embedded (FFPE) tissue was collected for tumour mutational signature profiling comprising:

1. 9 adenomas and 15 CRCs from 13 biallelic *MUTYH* cases;
2. 4 CRCs from 4 monoallelic *MUTYH* cases;
3. 7 adenomas, 1 hyperplastic polyp, 1 traditional serrated adenoma and 2 CRCs from 7 biallelic *NTHL1* cases and
4. 2 CRCs from 2 monoallelic *NTHL1* cases.

A reference/control group of 46 participants from the CCFR who developed mismatch repair (MMR)-proficient adenomas (n=27) and/or MMR-proficient CRCs (n=26) and who were confirmed to not carry LP/P variants in 16 hereditary CRC/polyposis genes as defined in Seifert *et al.* [8] (i.e., non-hereditary/sporadic cases) were included in this study.

The mutational signature profiles of 12 CRCs from eight biallelic *MUTYH* cases and 4 CRCs from 4 monoallelic *MUTYH* cases described above have been reported previously [5,6]. Two CRCs from two biallelic and two monoallelic *NTHL1* cases described above have been reported previously [7]. The studies were approved by the respective ethics committees and institutional review boards. All participants provided written informed consent for collection of tissue and peripheral blood samples.

### Whole-exome sequencing and bioinformatic analysis

Adenoma and CRC tissue DNA and matched blood-derived DNA underwent whole-exome sequencing (WES) using the SureSelect Clinical Research Exome v.2 kit (Agilent Technologies, Santa Clara, CA, United States), to a median depth of 357.9 reads (interquartile range (IQR)=287.8-464.0) for FFPE tissue DNA samples and median depth of 179.1 reads (IQR=118.1-204.6) for blood-derived DNA samples. Somatic single-nucleotide variants and short insertion/deletions were determined using the intersection of calls from Strelka (v.2.9.2) [9] and Mutect2 (v.4.0) [10]. Tumour mutation burden (TMB) was calculated as the total number of all somatic single-nucleotide variants and short insertion/deletions observed in a sample divided by the size of the capture region (67Mb). A threshold for including variants was chosen based on a minimum depth (50 reads) and a minimum variant allele frequency of 10% as previously published [5]. Mutational signature profiles were calculated using the simulated annealing method previously described by SignatureEstimation [11] using a reduced set of 16 SBS signatures (**Supplementary Table 1**) as previously determined to be present in the colon/colorectal cancer tissue [5–7,12–19]. The following RefSeq transcripts were used: NM_001128425.1 (*MUTYH*), NM_002528.7 (*NTHL1*), NM_001369786.1 (*KRAS*) and NM_006218.4 (*PIK3CA*).

### Statistical analysis

For each signature profile, we compared the biallelic cases with the corresponding CRCs or adenomas from the non-hereditary group. Statistical significance between two groups was determined using a two-sided *t-test* with a *p-value*<0.05 considered to be statistically significant. For group comparisons, one-way *ANOVA* was used. Additionally, we determined the *Cohen’s d* effect size to measure the difference between the means of two subgroups.

### Source code

All data analysis was performed using Python v.3.11 [20], Numpy v.1.24 [21] and Scikit-Learn v.1.3 [22]. Data visualisation was done using the R programming language v.4.3.2 [23] and RStudio v.0.16.0 [24] using the following packages: ggplot2 v.3.5.1 [25], cowplot v.1.1.3 [26] and dplyr v.1.1.4 [27].

## RESULTS

The clinicopathological characteristics of the participants and their adenomas and CRCs are shown in **Table 1**. The biallelic *MUTYH* and biallelic *NTHL1* cases are presented in **Supplementary Table 2**. Of note, all adenomas and CRCs were MMR-proficient by immunohistochemistry except for two biallelic *MUTYH* cases; Pat_301 (2xCRCs at 50 years, one MMR-proficient and one MMR-deficient with MLH1/PMS2 loss), and Pat_315 (1xCRC at 39 years with MSH2/MSH6 loss). The SBS mutational signature profiles of each adenoma and CRC included in the study are presented in **Figure 1**.

**Figure 1:**
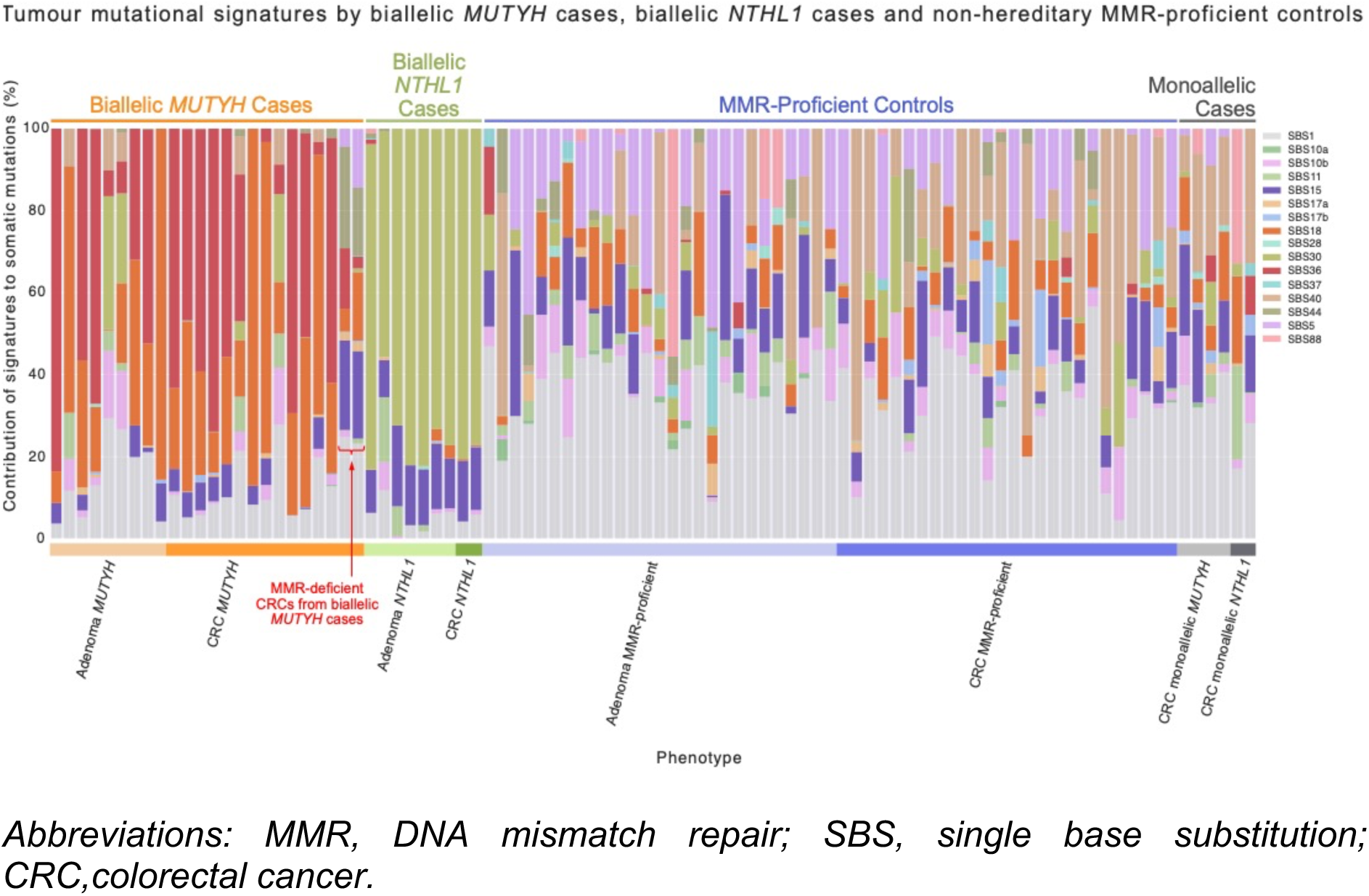
Mutational signatures observed across the cohort.

**Table 1.** The clinicopathological characteristics of the participants and their adenomas and CRCs from each of the biallelic *MUTYH* cases, biallelic *NTHL1* cases and the adenomas and CRCs from the non-hereditary (control) groups included in this study. Overview of the phenotypes by sex, age at diagnosis (including mean and standard deviation), anatomical site, histological type, T stage, grade of tumour and study separated by adenoma and colorectal cancer tissue type and case subgroups.

|  | *MUTYH* cases (n=13) |  | *NTHL1* cases (n=5) |  | Non-hereditary Controls (n=46) |  | Total (n=64) Individuals |
| --- | --- | --- | --- | --- | --- | --- | --- |
|  | Adenoma biallelic *MUTYH* (n=9, 10.5%) | CRC biallelic *MUTYH* (n=15, 17.4%) | Adenoma biallelic *NTHL1* (n=7, 8.1%) | CRC biallelic *NTHL1* (n=2, 2.3%) | MMR-proficient Adenomas (n=27, 31.4%) | MMR-proficient CRCs (n=26, 30.2%) | Total (n=86, 100%) Tissues |
| Sex, n (%) |  |  |  |  |  |  |  |
| Male | 5 (55.6%) | 11 (73.3%) | 3 (42.9%) | 0 (0.0%) | 12 (44.4%) | 12 (46.2%) | 43 (50.0%) |
| Female | 4 (44.4%) | 4 (26.7%) | 4 (57.1%) | 2 (100.0%) | 15 (55.6%) | 14 (53.8%) | 43 (50.0%) |
| Age at diagnosis, n (%) |  |  |  |  |  |  |  |
| Mean ± SD | 52.3 ± 14.4 | 52.2 ± 11.1 | 57.7 ± 5.7 | 68.5 ± 10.6 | 43.7 ± 10.6 | 42.8 ± 9.1 | 48.0 ± 12.1 |
| Min. - Max. | 33 - 73 | 33 - 64 | 51 - 66 | 61 - 76 | 27 - 61 | 21 - 59 | 21 - 76 |
| $\leq 50$ years | 2 (22.2%) | 6 (40.0%) | 0 (0.0%) | 0 (0.0%) | 19 (70.4%) | 22 (84.6%) | 49 (57.0%) |
| $> 50$ years | 7 (77.8%) | 9 (60.0%) | 7 (100.0%) | 2 (100.0%) | 8 (29.6%) | 4 (15.4%) | 37 (43.0%) |
| Ethnicity, n (%) |  |  |  |  |  |  |  |
| European | 9<br>(100.0%) | 15<br>(100.0%) | 7<br>(100.0%) | 2<br>(100.0%) | 27<br>(100.0%) | 24<br>(92.3%) | 84<br>(97.7%) |
| East Asian | 0 (0.0%) | 0 (0.0%) | 0 (0.0%) | 0 (0.0%) | 0 (0.0%) | 1 (3.8%) | 1 (1.2%) |
| South Asian | 0 (0.0%) | 0 (0.0%) | 0 (0.0%) | 0 (0.0%) | 0 (0.0%) | 1 (3.8%) | 1 (1.2%) |
| Anatomical site, n (%) |  |  |  |  |  |  |  |
| Proximal colon | 3<br>(33.3%) | 12<br>(80.0%) | 2<br>(28.6%) | 2<br>(100.0%) | 9 (33.3%) | 12<br>(46.2%) | 40<br>(46.5%) |
| Distal colon | 0 (0.0%) | 1 (6.7%) | 0 (0.0%) | 0 (0.0%) | 6 (22.2%) | 8<br>(30.8%) | 15<br>(17.4%) |
| Rectum | 0 (0.0%) | 2<br>(13.3%) | 1<br>(14.3%) | 0 (0.0%) | 6 (22.2%) | 6<br>(23.1%) | 15<br>(17.4%) |
| Unknown | 6<br>(66.7%) | 0 (0.0%) | 4<br>(57.1%) | 0 (0.0%) | 6 (22.2%) | 0 (0.0%) | 16<br>(18.6%) |
| Colorectal Adenoma Histological Type, n (%) |  |  |  |  |  |  |  |
| Tubular adenoma | 2<br>(22.2%) | - | 5<br>(71.4%) | - | 14<br>(51.9%) | - | - |
| Tubulovillous adenoma | 4<br>(44.4%) | - | 2<br>(28.6%) | - | 13<br>(48.1%) | - | - |
| Unknown | 3<br>(33.3%) | - | 0 (0.0%) | - | 0 (0.0%) | - | - |
| CRC Histological Type, n (%) |  |  |  |  |  |  |  |
| Adenocarcinoma | - | 15<br>(100.0%) | - | 2<br>(100.0%) | - | 24<br>(92.3%) | - |
| Mucinous adenocarcinoma | - | 0 (0.0%) | - | 0 (0.0%) | - | 0 (0.0%) | - |
| Signet ring adenocarcinoma | - | 0 (0.0%) | - | 0 (0.0%) | - | 1 (3.8%) | - |
| Undifferentiated (incl. medullary) | - | 0 (0.0%) | - | 0 (0.0%) | - | 1 (3.8%) | - |
| Grade of CRC, n (%) |  |  |  |  |  |  |  |
| Well differentiated | - | 1 (6.7%) | - | 0 (0.0%) | - | 3<br>(11.5%) | - |
| Moderately differentiated | - | 13<br>(86.7%) | - | 2<br>(100.0%) | - | 16<br>(61.5%) | - |
| Poorly differentiated | - | 0 (0.0%) | - | 0 (0.0%) | - | 4<br>(15.4%) | - |
| Unknown | - | 1 (6.7%) | - | 0 (0.0%) | - | 3<br>(11.5%) | - |
| Study, n (%) |  |  |  |  |  |  |  |
| ANGELS | 0 (0.0%) | 0 (0.0%) | 4<br>(57.1%) | 0 (0.0%) | 0 (0.0%) | 0 (0.0%) | 4 (4.7%) |
| CCFR | 6<br>(66.7%) | 14<br>(93.3%) | 0 (0.0%) | 1<br>(50.0%) | 27<br>(100.0%) | 27<br>(103.8%) | 74<br>(86.0%) |
| GCPS | 3<br>(33.3%) | 1 (6.7%) | 3<br>(42.9%) | 1<br>(50.0%) | 0 (0.0%) | 0 (0.0%) | 8 (9.3%) |

### The SBS18+SBS36 mutational signature is elevated in both adenomas and CRCs from biallelic MUTYH cases

The mean (±standard deviation) proportion of SBS18+SBS36 in the adenomas (65.6%±29.6%) and MMR-proficient CRCs (76.2%±20.5%) from biallelic *MUTYH* cases were not significantly different (*p-value*=0.37) (**Figure 2A**, **Table 2**). This result is further highlighted when comparing the SBS18+SB36 proportions in adenomas and CRCs from the same participant (**Figure 3**). In contrast, the mean proportion of SBS18+SBS36 in adenomas and CRCs from biallelic *MUTYH* cases were significantly higher compared with the mean proportion in non-hereditary adenomas (65.6%±29.6% versus 7.6%±7.0%, *p-value*=3.4x10^-4^) and CRCs (76.2%±20.5% versus 6.5%±5.5%, *p-value*=2.2x10^-8^) (**Figure 2A**, **Table 3**).

**Figure 2:**
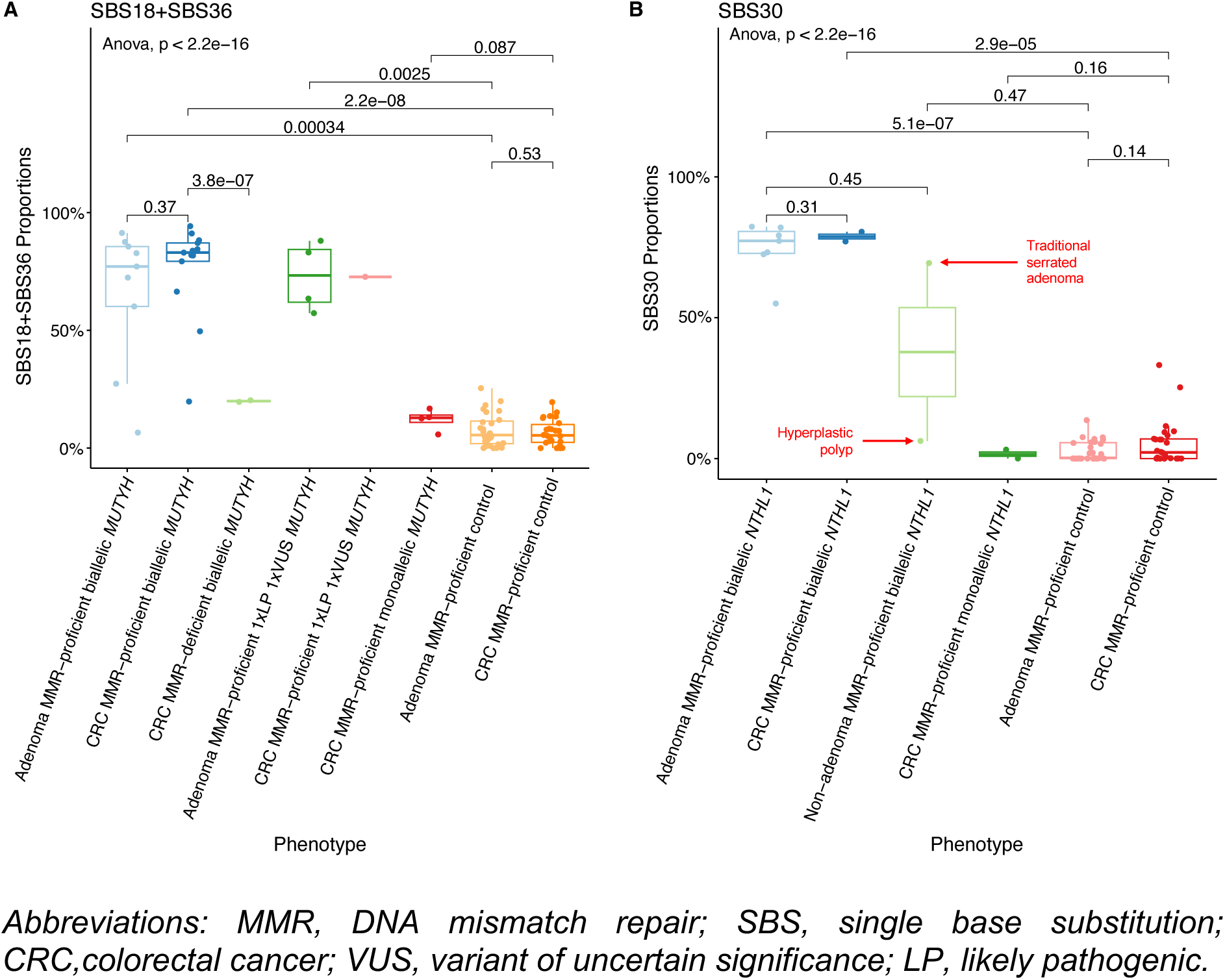
Boxplots of whole-exome sequencing derived genomic features for A) SBS18+SBS36 proportions in *MUTYH* cases and non-hereditary groups and B) SBS30 proportions in *NTHL1* cases and non-hereditary groups.

**Figure 3.**
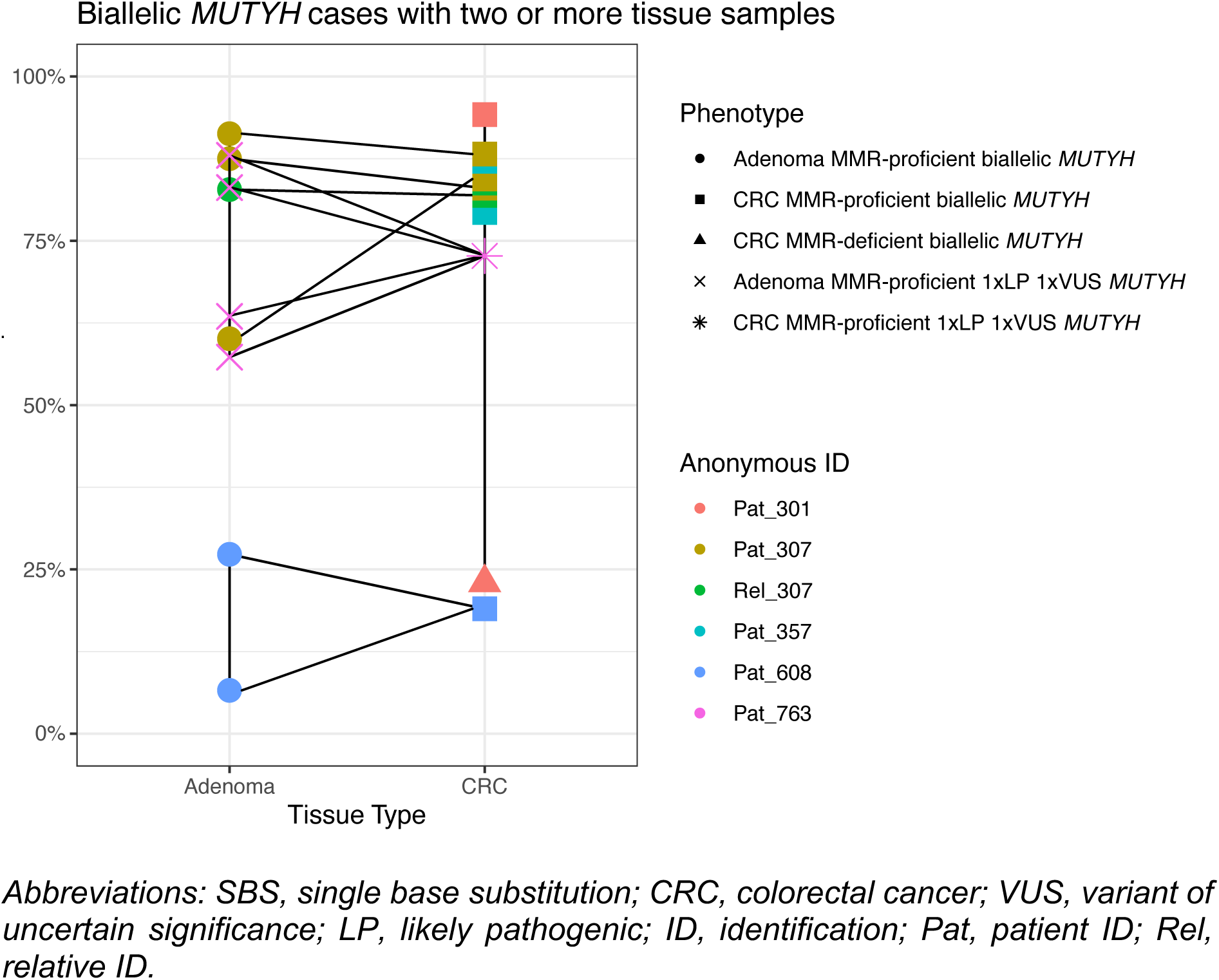
Line plot displaying the comparison of SBS18+SBS36 signature proportions for adenomas and colorectal cancers related to each biallelic *MUTYH* case and for the participant with a pathogenic and variant of uncertain significance in *MUTYH* (Pat_763).

**Table 2.** The mean, standard deviation, and range of five genomic features derived from whole-exome sequencing testing for their differences between tissue type and by *MUTYH* or *NTHL1* case or non-hereditary status. Statistically significant p-values are highlighted in **bold**.

|  | Biallelic cases *MUTYH* |  |  |  | Biallelic cases *NTHL1* |  |  |  | Proficient Controls |  |  |  |  |
| --- | --- | --- | --- | --- | --- | --- | --- | --- | --- | --- | --- | --- | --- |
|  | Adenom<br>a biallelic<br>*MUTYH*<br>(n=9,<br>10.7%) | CRC<br>biallelic<br>*MUTYH*<br>(n=13,<br>15.5%)<sup>1</sup> | t-test<br>(p-value)<br><sup>2</sup> | Effect<br>Size<br>(Cohen'<br>s d) | Adenom<br>a biallelic<br>*NTHL1*<br>(n=7,<br>8.3%) | CRC<br>biallelic<br>*NTHL1*<br>(n=2,<br>2.4%) | t-test<br>(p-value)<br><sup>2</sup> | Effect<br>Size<br>(Cohen'<br>s d) | Adenom<br>a<br>proficient<br>control<br>(n=27,<br>32.1%) | CRC<br>proficient<br>control<br>(n=26,<br>31.0%) | t-test<br>(p-value)<br><sup>2</sup> | Effect<br>Size<br>(Cohen'<br>s d) | Total<br>(n=84,<br>100%) |
| SBS18+SBS<br>36 |  |  | 0.37 | -0.4 |  |  | 0.30 | 0.5 |  |  | 0.53 | 0.2 |  |
| Mean | 65.6% | 76.2% |  |  | 1.0% | 0.4% |  |  | 7.6% | 6.5% |  |  | 23.4% |
| SD | 29.6% | 20.5% |  |  | 1.4% | 0.0% |  |  | 7.0% | 5.5% |  |  | 32.0% |
| Range | 6.6% -<br>91.3% | 19.8% -<br>94.2% |  |  | 0% -<br>3.2% | 0.4% -<br>0.4% |  |  | 0% -<br>25.4% | 0% -<br>19.5% |  |  | 0% -<br>94.2% |
| SBS30 |  |  | 0.38 | 0.4 |  |  | 0.31 | -0.5 |  |  | 0.14 | -0.4 |  |
| Mean | 6.0% | 2.0% |  |  | 74.5% | 78.8% |  |  | 2.8% | 5.4% |  |  | 11.6% |
| SD | 12.3% | 6.0% |  |  | 9.4% | 2.4% |  |  | 3.6% | 8.0% |  |  | 23.4% |
| Range | 0% -<br>32.4% | 0% -<br>21.4% |  |  | 55% -<br>82.3% | 77.1% -<br>80.5% |  |  | 0% -<br>13.6% | 0% -<br>33.2% |  |  | 0% -<br>82.3% |
| TMB |  |  | 0.12 | -0.8 |  |  | 0.87 | -0.1 |  |  | **7.4x10<sup>-5</sup>** | -1.2 |  |
| Mean | 4.8 | 7.5 |  |  | 7.1 | 7.5 |  |  | 1.5 | 2.8 |  |  | 3.8 |
| SD | 4.3 | 2.9 |  |  | 4.2 | 2.0 |  |  | 0.9 | 1.3 |  |  | 3.3 |
| Range | 0.3 - 12.7 | 3.5 - 13.3 |  |  | 2.5 -<br>14.8 | 6.1 - 8.8 |  |  | 0.3 - 4.1 | 1.4 - 5.9 |  |  | 0.3 -<br>14.8 |
| INDEL count |  |  | **0.02** | -1.1 |  |  | 0.95 | 0.0 |  |  | **2.3x10<sup>-3</sup>** | -0.9 |  |
| Mean | 4.4 | 9.7 |  |  | 10.7 | 10.5 |  |  | 7.1 | 11.3 |  |  | 8.9 |
| SD | 4.9 | 4.6 |  |  | 9.3 | 0.7 |  |  | 4.2 | 5.4 |  |  | 5.6 |
| Range | 1 - 17 | 2 - 16 |  |  | 2 - 25 | 10 - 11 |  |  | 1 - 17 | 3 - 20 |  |  | 1 - 25 |
| SNV count |  |  | 0.13 | -0.7 |  |  | 0.87 | -0.1 |  |  | **9.8x10<sup>-5</sup>** | -1.2 |  |
| Mean | 318.6 | 494.3 |  |  | 466.9 | 491.0 |  |  | 91.1 | 178.1 |  |  | 245.6 |
| SD | 285.4 | 197.5 |  |  | 274.5 | 132.9 |  |  | 56.8 | 86.9 |  |  | 217.9 |
| Range | 19 - 851 | 231 - 895 |  |  | 168 - 974 | 397 - 585 |  |  | 16 - 262 | 84 - 381 |  |  | 16 - 974 |

*Abbreviations: CRC, colorectal cancer; SBS, single base substitution; SD, standard deviation; TMB, tumour mutation burden; INDEL, large insertion/deletion; SNV, single nucleotide variant.*
<sup>1</sup> MMR-proficient CRCs only from biallelic *MUTYH* cases were included.
<sup>2</sup> two-tailed t-test.

**Table 3.** The mean, standard deviation, and range of five genomic features derived from whole-exome sequencing testing assessed for their differences between *MUTYH* or *NTHL1* case or non-hereditary status for colorectal adenomas and colorectal cancers separately. Statistically significant p-values are highlighted in **bold**.

|  | Colorectal Adenomas |  |  |  | Colorectal Cancers |  |  |  | Colorectal Adenomas |  |  |  | Colorectal Cancers |  |  |  |  |
| --- | --- | --- | --- | --- | --- | --- | --- | --- | --- | --- | --- | --- | --- | --- | --- | --- | --- |
|  | Biallelic MUTYH cases (n=9, 10.7%) | Non-hereditary controls (n=27, 32.1%) | t-test (p-value)<sup>2</sup> | Effect Size (Cohen's d) | Biallelic MUTYH cases (n=13, 15.5%) | Non-hereditary controls (n=26, 31.0%) | t-test (p-value)<sup>2</sup> | Effect Size (Cohen's d) | Biallelic NTHL1 cases (n=7, 8.3%) | Non-hereditary controls (n=27, 32.1%) | t-test (p-value)<sup>2</sup> | Effect Size (Cohen's d) | Biallelic NTHL1 cases (n=2, 2.4%) | Non-hereditary controls (n=26, 31.0%) | t-test (p-value)<sup>2</sup> | Effect Size (Cohen's d) | Total (n=84, 100%) |
| SBS18+SBS36 |  |  | **3.4x 10<sup>-4</sup>** | 3.7 |  |  | **2.2x 10<sup>-8</sup>** | 5.6 |  |  | **8.0x 10<sup>-5</sup>** | -1.0 |  |  | **6.4x 10<sup>-6</sup>** | -1.1 |  |
| Mean | 65.6% | 7.6% |  |  | 76.2% | 6.5% |  |  | 1.0% | 7.6% |  |  | 0.4% | 6.5% |  |  | 23.4% |
| SD | 29.6% | 7.0% |  |  | 20.5% | 5.5% |  |  | 1.4% | 7.0% |  |  | 0.0% | 5.5% |  |  | 32.0% |
| Range | 6.6% - 91.3% | 0% - 25.4% |  |  | 19.8% - 94.2% | 0% - 19.5% |  |  | 0% - 3.2% | 0% - 25.4% |  |  | 0.4% - 0.4% | 0% - 19.5% |  |  | 0% - 94.2% |
| SBS30 |  |  | 0.45 | 0.5 |  |  | 0.15 | -0.5 |  |  | **5.1x 10<sup>-7</sup>** | 13.8 |  |  | **2.9x 10<sup>-5</sup>** | 9.3 |  |
| Mean | 6.0% | 2.8% |  |  | 2.0% | 5.4% |  |  | 74.5% | 2.8% |  |  | 78.8% | 5.4% |  |  | 11.6% |
| SD | 12.3% | 3.6% |  |  | 6.0% | 8.0% |  |  | 9.4% | 3.6% |  |  | 2.4% | 8.0% |  |  | 23.4% |
| Range | 0% - 32.4% | 0% - 13.6% |  |  | 0% - 21.4% | 0% - 33.2% |  |  | 55% - 82.3% | 0% - 13.6% |  |  | 77.1% - 80.5% | 0% - 33.2% |  |  | 0% - 82.3% |
| TMB |  |  | **4.7x**<br>**10<sup>-2</sup>** | 1.5 |  |  | **7.5x**<br>**10<sup>-5</sup>** | 2.3 |  |  | **1.2x**<br>**10<sup>-2</sup>** | 2.8 |  |  | 0.18 | 3.4 |  |
| Mean | 4.8 | 1.5 |  |  | 7.5 | 2.8 |  |  | 7.1 | 1.5 |  |  | 7.5 | 2.8 |  |  | 3.8 |
| SD | 4.3 | 0.9 |  |  | 2.9 | 1.3 |  |  | 4.2 | 0.9 |  |  | 2.0 | 1.3 |  |  | 3.3 |
| Range | 0.3 -<br>12.7 | 0.3 -<br>4.1 |  |  | 3.5 -<br>13.3 | 1.4 -<br>5.9 |  |  | 2.5 -<br>14.8 | 0.3 -<br>4.1 |  |  | 6.1 -<br>8.8 | 1.4 -<br>5.9 |  |  | 0.3 -<br>14.8 |
| INDEL<br>count |  |  | 0.18 | -0.6 |  |  | 0.33 | -0.3 |  |  | 0.35 | 0.7 |  |  | 0.48 | -0.2 |  |
| Mean | 4.4 | 7.1 |  |  | 9.7 | 11.3 |  |  | 10.7 | 7.1 |  |  | 10.5 | 11.3 |  |  | 8.9 |
| SD | 4.9 | 4.2 |  |  | 4.6 | 5.4 |  |  | 9.3 | 4.2 |  |  | 0.7 | 5.4 |  |  | 5.6 |
| Range | 1 - 17 | 1 - 17 |  |  | 2 - 16 | 3 - 20 |  |  | 2 -<br>25 | 1 - 17 |  |  | 10 -<br>11 | 3 - 20 |  |  | 1 -<br>25 |
| SNV count |  |  | **4.4x**<br>**10<sup>-2</sup>** | 1.5 |  |  | **6.9x**<br>**10<sup>-5</sup>** | 2.4 |  |  | **1.1x**<br>**10<sup>-2</sup>** | 2.9 |  |  | 0.18 | 3.5 |  |
| Mean | 318.6 | 91.1 |  |  | 494.3 | 178.1 |  |  | 466.<br>9 | 91.1 |  |  | 491.<br>0 | 178.1 |  |  | 245.<br>6 |
| SD | 285.4 | 56.8 |  |  | 197.5 | 86.9 |  |  | 274.<br>5 | 56.8 |  |  | 132.<br>9 | 86.9 |  |  | 217.<br>9 |
| Range | 19 -<br>851 | 16 -<br>262 |  |  | 231 -<br>895 | 84 -<br>381 |  |  | 168 -<br>974 | 16 -<br>262 |  |  | 397 -<br>585 | 84 -<br>381 |  |  | 16 -<br>974 |

Abbreviations: CRC, colorectal cancer; SBS, single base substitution; SD, standard deviation; TMB, tumour mutation burden; INDEL, large insertion/deletion; SNV, single nucleotide variant.
<sup>1</sup> MMR-proficient CRCs only from biallelic *MUTYH* cases were included
<sup>2</sup> Two-tailed t-test

### Co-occurrence of mutational processes related to defective MUTYH and defective DNA mismatch repair

In the two MMR-deficient CRCs from biallelic *MUTYH* cases (Pat_301 and Pat_315), the mean proportion of SBS18+SBS36, was significantly lower compared with the MMR-proficient CRCs from biallelic *MUTYH* cases (20.0%±0.5% versus 76.2%±20.5%, *p-value*=3.8x10^-7^) but they were still higher compared with non-hereditary CRCs (6.5%±5.5%, *p-value*=2.2x10^-8^) (**Figure 2A**, **Table 3**). Both these MMR-deficient CRCs also showed higher proportions of SBS15 and SBS44, which are mutational signatures associated with MMR-deficiency (**Figure 1**). In addition, the TMB of these two MMR-deficient CRCs (53.9 and 25.4 mutations/Mb, respectively) was higher compared with the mean TMB of the MMR-proficient CRCs from biallelic *MUTYH* cases (7.5±2.9 mutations/Mb) (**Figure 4**).

The cause of MMR-deficiency in Pat_301 and Pat_315 was not related to carrying a germline LP/P variant in one of the DNA MMR genes, but rather from two somatic MMR mutations causing biallelic inactivation in each CRC as determined from the WES data. The CRC showing loss of MLH1/PMS2 protein expression from Pat_301 had two somatic mutations in *MLH1* (c.1813G>T p.(Glu605Ter) and c.1816G>T p.(Gly606Ter)) and no evidence of tumour *MLH1* promoter hypermethylation. The CRC showing loss of MSH2/MSH6 protein expression from Pat_315 had a somatic mutation in *MSH2* (c.394G>T p.(Glu132Ter)) and loss of heterozygosity indicating loss of the wildtype *MSH2* allele. The somatic single nucleotide mutations observed in *MLH1* and *MSH2* matched the mutational contexts associated with SBS18 and SBS36 (Pat_301:TCT>TAT and TCC>TAC; Pat_315:TCA>TTA), suggesting the constitutionally defective *MUTYH* contributed to these somatic MMR mutational events and resulted in MMR-deficiency in these two CRCs. Interestingly, the synchronous MMR-proficient CRC from Pat_301 exhibited a high proportion of SBS18+SBS36 (94.2%) and a low TMB (9.4 mutations/Mb), further highlighting the impact of tumour MMR-deficiency on the SBS18+SBS36 signature proportions in biallelic *MUTYH* cases.

### Somatic mutations as biomarkers of biallelic MUTYH status in adenomas

Previously, the *KRAS* c.34G>T p.(Gly12Cys) and *PIK3CA* c.1636C>A p.(Gln546Lys) somatic mutations were shown to be recurrent mutations significantly increased in CRCs from biallelic *MUTYH* pathogenic variant cases [6]. In adenomas, the *KRAS* c.34G>T mutation was present in 6/9 (66.7%) and 2/27 (7.4%) of the biallelic *MUTYH* and non-hereditary adenomas, respectively (*p-value*=3.1x10^-2^). The *KRAS* c.34G>T mutation had a positive predictive value of 75% and a negative predictive value of 89.3% in adenomas compared with a positive predictive value of 100% and negative predictive value of 86.7% in CRCs, indicating that the somatic *KRAS* mutation may not be as clinically useful in identifying biallelic *MUTYH* cases in adenomas as it is in CRCs. The *PIK3CA* c.1636C>A mutation was not observed in adenomas from biallelic *MUTYH* cases or in adenomas from the non-hereditary group (**Supplementary Figure 1**).

### The SBS18+SBS36 mutational signature provides evidence for variant classification

We profiled four adenomas and a CRC from Pat_763 who carried a germline heterozygous pathogenic variant (c.1187G>A p.(Gly396Asp)) and a germline heterozygous VUS (c.533G>C p.(Gly178Ala)) in *MUTYH.* All four adenomas (mean proportion: 73.0%±14.9%, range: 57.3%-88.0%) and the CRC (72.7%) demonstrated high proportions of SBS18+SBS36 consistent with germline biallelic inactivation of *MUTYH* (**Figure 2A**, **Figure 3**). No somatic second hits in *MUTYH* were observed that may have accounted for the high SBS18+SBS36 signature proportions in the adenomas and CRC. These findings support a reclassification of the *MUTYH* c.533G>C p.(Gly178Ala) variant as likely pathogenic.

### The SBS30 mutational signature is elevated in both adenomas and CRCs from biallelic NTHL1 cases

The mean proportion of SBS30 in adenomas (74.5%±9.4%) and CRCs (78.8%±2.4%) from biallelic *NTHL1* cases were not significantly different (*p-value*=0.31) (**Figure 2B**, **Table 2**). The mean proportion of SBS30 in adenomas from biallelic *NTHL1* cases was, however, significantly higher compared with the mean proportion in non-hereditary adenomas (74.5%±9.4% versus 2.8%±1.3%; p-value=5.1x10^-7^) (**Figure 2B**, **Table 3**). In addition to 7 adenomas and 2 CRCs, a hyperplastic polyp and a traditional serrated adenoma from two biallelic *NTHL1* cases (Pat_005 and Pat_469) were tested. Of note, the traditional serrated adenoma showed high proportion of SBS30 at 69.4%, whereas the SBS30 proportion in the hyperplastic polyp was only 6.2% (**Figure 2B**).

## DISCUSSION

In this study, we showed that the SBS18+SBS36 and SBS30 mutational signatures associated with biallelic *MUTYH* and biallelic *NTHL1* deficiencies, were present in adenomas at similar proportions to those observed in CRCs and were significantly higher when compared with the proportions observed in non-hereditary adenomas and CRCs. Together, these results demonstrate the presence of these mutational processes and consequent mutational signatures, at diagnostic levels in the pre-malignant stage, thereby enabling the opportunity for early CRC detection by expanding the potential tissue available for profiling.

We identified two scenarios where SBS30 or SBS18+SBS36 may present with limitations. Firstly, although SBS30 was shown to be a predominant mutational signature in adenomas from biallelic *NTHL1* cases, our results showed variable presence of SBS30 in two serrated polyp subtypes, 69.4% in the traditional serrated adenoma and only 6.2% in the hyperplastic polyp. As biallelic *NTHL1* cases can present with mixed polyp types [28], further research is needed to determine the utility of testing serrated polyps for mutational signatures for *NTHL1* and more broadly for other hereditary CRC/polyposis syndromes. Secondly, we tested two MMR-deficient CRCs from two biallelic *MUTYH* cases where the mutational signature profile showed defective MMR related to the presence of SBS15 and SBS44 and a hypermutated TMB that co-occurred with the SBS18+SBS36 signature, albeit at lower proportions than observed in MMR-proficient CRCs from biallelic *MUTYH* cases. These findings highlight MMR-deficiency as an important diagnostic caveat for utilising SBS18+SBS36 to identify biallelic *MUTYH* cases or for classifying variants.

This study extends on our previous work for applying SBS18+SBS36 in CRCs to reclassify VUSs in *MUTYH* [5]. We showed high levels of SBS18+SBS36 in the CRC and multiple adenomas from the same person provides high confidence that the *MUTYH* c.533G>C p.(Gly178Ala) variant is pathogenic. Additional evidence related to its absence in gnomAD and from *in-silico* predictions from REVEL, SIFT, PolyPhen-2 and Align-GVGD suggest this missense change affects protein function, further supporting pathogenicity (https://www.ncbi.nlm.nih.gov/clinvar/variation/481808/, last accessed date: August 1^st^, 2024). The ability to test multiple independent adenomas/CRCs provides high confidence for variant classification where all or none of the lesions have the signature. The clinical genetics community is increasingly challenged by VUS, where around half (47.8%, 1329/2782) of *MUTYH* variants in ClinVar are currently classified as VUS (stand: August 6^th^, 2024) [29]. Approaches to classify variants with existing and widely used infrastructure i.e., next generation sequencing and validated bioinformatic tools, will aid in reclassifying variants and optimising clinical management and cancer prevention for the patient and their relatives.

Limitations of this study include the lack of ethnic diversity within the case and non-hereditary groups which were predominantly white European. Similarly, there was a limited range of germline LP/P variants for both *MUTYH* and *NTHL1*. The consistency of mutational signature findings across a broader group of cases of different pathogenic variants and ethnic backgrounds would provide evidence of the robustness of this approach. All of the CRCs and adenomas tested in this study were from FFPE tissue, however we have previously shown that mutational signature profiling is effective in both FFPE and fresh frozen tissue DNA samples [5].

## CONCLUSIONS

This study provides important findings demonstrating that testing adenomas for SBS18+SBS36 or SBS30 can be an equally effective alternative to identifying biallelic *MUTYH* or biallelic *NTHL1* cases, respectively, if CRC has not yet developed or tissue is not available. This provides important opportunities for clinical management decision-making such as colectomy versus endoscopic polypectomy for CRC prevention given the established high CRC penetrance in biallelic cases. Furthermore, the specificity of these signatures enables the utility of mutational signature profiling to classify VUS. Our study identified potential caveats to using mutational signatures diagnostically, namely, the presence of MMR-deficiency which may diminish the SBS18+SBS36 signature, while for SBS30, testing of serrated polyps needs further investigation. This study adds to the growing evidence of the clinical utility of gene specific mutational signature profiling for identifying hereditary CRC/polyposis syndromes and further expands the opportunities to utilise mutational signatures as a supportive feature for variant classification.

## Data Availability

The datasets used and/or analysed during the current study are available from the corresponding author on reasonable request.

## ACKNOWLEDGEMENTS

The authors thank members of the Colorectal Oncogenomics Group and members from the Genomic Medicine and Family Cancer Clinic for their support of this manuscript. We thank the participants and staff from the ANGELS study, GCPS study, and from the Australasian and Ontario sites of the CCFR, in particular Maggie Angelakos, Samantha Fox, Cary Greenberg and Allyson Templeton for their support of this manuscript. The CCFR graciously thanks the generous contributions of their study participants, dedication of study staff, and the financial support from the U.S. National Cancer Institute, without which this important registry would not exist. The authors thank Melbourne Bioinformatics and the Australian Genome Research Facility for their collaboration on this project.

## AUTHOR CONTRIBUTIONS

**Romy Walker:** Conceptualization, Methodology, Software, Validation, Formal analysis, Investigation, Data Curation, Writing - Original Draft, Writing - Review & Editing, Visualization, Funding acquisition. **Jihoon E. Joo:** Software, Formal analysis, Investigation, Writing - Review & Editing, Visualization. **Khalid Mahmood:** Software, Investigation, Writing - Review & Editing. **Mark Clendenning:** Investigation, Writing - Review & Editing. **Julia Como:** Investigation, Writing - Review & Editing. **Susan G. Preston:** Investigation, Writing - Review & Editing. **Sharelle Joseland:** Resources, Writing - Review & Editing. **Bernard J. Pope:** Software, Writing - Review & Editing, Funding acquisition. **Ana B. D. Medeiros:** Formal analysis, Resources, Writing - Review & Editing. **Brenely V. Murillo:** Resources, Writing - Review & Editing. **Nicholas Pachter:** Resources, Writing - Review & Editing. **Kevin Sweet:** Resources, Writing - Review & Editing. **Allan D. Spigelman:** Resources, Writing - Review & Editing. **Alexandra Groves:** Resources, Writing - Review & Editing. **Margaret Gleeson:** Resources, Writing - Review & Editing. **Krzysztof Bernatowicz:** Resources, Writing - Review & Editing. **Nicola Poplawski:** Resources, Writing - Review & Editing. **Lesley Andrews:** Resources, Writing - Review & Editing. **Emma Healey:** Resources, Writing - Review & Editing. **Steven Gallinger:** Resources, Writing - Review & Editing. **Robert C. Grant:** Resources, Writing - Review & Editing. **Aung K. Win:** Resources, Writing - Review & Editing, Funding acquisition. **John L. Hopper:** Resources, Writing - Review & Editing, Funding acquisition. **Mark A. Jenkins:** Resources, Writing - Review & Editing, Funding acquisition. **Giovana T. Torrezan:** Conceptualization, Resources, Data Curation, Writing - Review & Editing. **Christophe Rosty:** Data Curation, Writing - Review & Editing. **Finlay A. Macrae:** Resources, Writing - Review & Editing. **Ingrid M. Winship:** Resources, Writing - Review & Editing. **Daniel D. Buchanan:** Conceptualization, Methodology, Validation, Data Curation, Investigation, Resources, Writing - Review & Editing, Supervision, Project administration, Funding acquisition. **Peter Georgeson:** Conceptualization, Methodology, Software, Validation, Data Curation, Formal analysis, Investigation, Writing - Review & Editing, Visualization, Supervision, Funding acquisition.

