## Supplementary Material for "Adenomas from individuals with pathogenic biallelic variants in the *MUTYH* and *NTHL1* genes demonstrate base excision repair tumour mutational signature profiles similar to colorectal cancers, expanding potential diagnostic and variant classification applications"

### SUPPLEMENTARY FIGURE

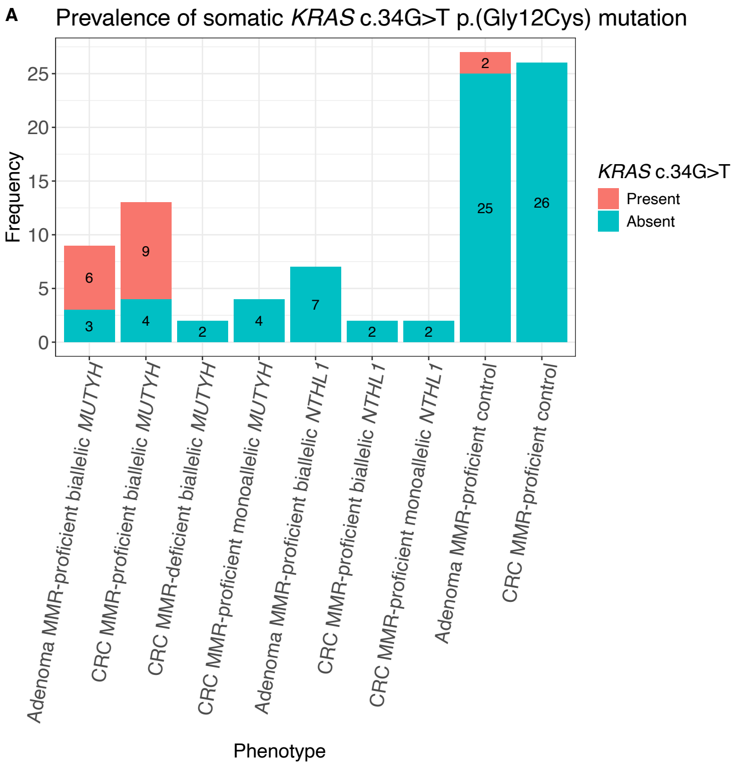

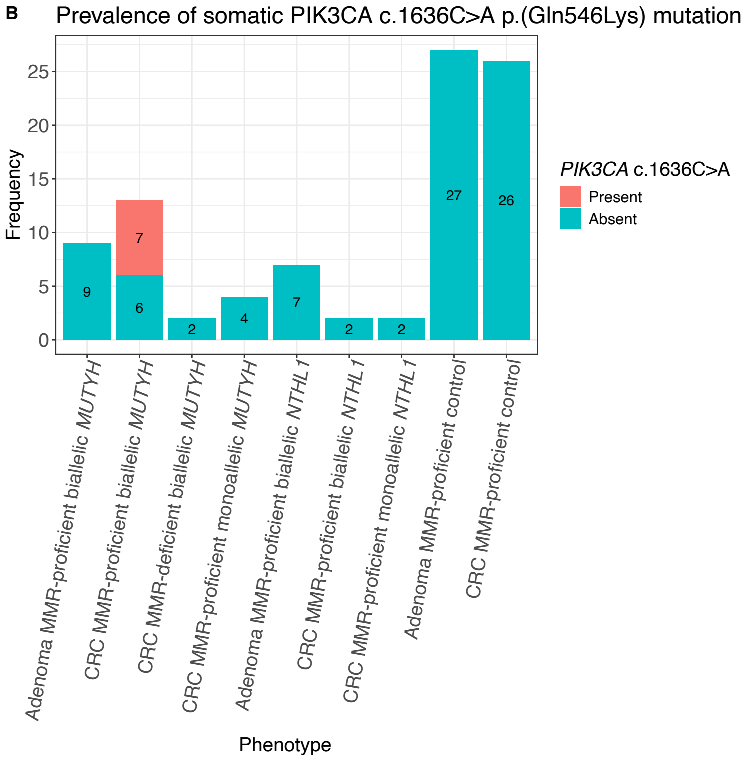

**Supplementary Figure S1.** Bar plots displaying the proportion of known hotspot mutations A) *KRAS* c.34G>T p.(Gly12Cys) and B) *PIK3CA* c.1636C>A p.(Gln546Lys) in the adenomas and CRCs in this study. *Abbreviations: MMR, DNA mismatch repair; CRC, colorectal cancer*.

### SUPPLEMENTARY TABLES

#### **Supplementary Table S1.** Reduced set of mutational signatures used in this study and their proposed aetiologies (if known).

| # | Signature | Proposed Aetiology | Reason for inclusion | Reference[1] |
|---|---|---|---|---|
| 1 | SBS1 | Spontaneous deamination of 5-methylcytosine (clock-like signature) | Set of signatures observed in 59 whole-exome sequenced CRCs | Alexandrov *et al.* , 2020; Everall *et al.* , 2023 |
| 2 | SBS5 | Unknown (clock-like signature) | Set of signatures observed in 59 whole-exome sequenced CRCs | Alexandrov *et al.* , 2020 |
| 3 | SBS10a | Polymerase epsilon exonuclease domain mutations | Set of signatures observed in 59 whole-exome sequenced CRCs | Alexandrov *et al.* , 2020; Everall *et al.* , 2023 |
| 4 | SBS10b | Polymerase epsilon exonuclease domain mutations | Set of signatures observed in 59 whole-exome sequenced CRCs | Alexandrov *et al.* , 2020; Everall *et al.* , 2023 |
| 5 | SBS11 | Red meat intake; Temozolomide treatment | Observed in indiviuals with a high unprocessed red meat consumption (high-risk for causing CRC) and observed in CRCs that were treated with Temozolomide (immune-checkpoint inhibitor blockade) | Gurjao *et al.* , 2021; Crisafulli *et al.* , 2022 |
| 6 | SBS15 | Defective DNA mismatch repair | Set of signatures observed in 59 whole-exome sequenced CRCs | Alexandrov *et al.* , 2020; Everall *et al.* , 2023 |
| 7 | SBS17a | Unknown | Set of signatures observed in 59 whole-exome sequenced CRCs | Alexandrov *et al.* , 2020; Everall *et al.* , 2023 |
| 8 | SBS17b | Unknown | Set of signatures observed in 59 whole-exome sequenced CRCs | Alexandrov *et al.* , 2020; Everall *et al.* , 2023 |
| 9 | SBS18 | Damage by reactive oxygen species | Set of signatures observed in 59 whole-exome sequenced CRCs | Alexandrov *et al.* , 2020; Everall *et al.* , 2023 |
| 10 | SBS28 | Unknown | Set of signatures observed in 59 whole-exome sequenced CRCs | Alexandrov *et al.* , 2020; Everall *et al.* , 2023 |
| 11 | SBS30 | Defective base excision repair due to *NTHL1* mutations | Observed in CRC-affected individuals diagnosed with *NTHL1* -associated polyposis | Grolleman *et al.* , 2019 |
| 12 | SBS36 | Defective base excision repair due to *MUTYH* mutation | Observed in CRC-affected individuals diagnosed with *MUTYH* -associated polyposis | Viel *et al.* , 2017, Georgeson *et al.* , 2022 |
| 13 | SBS37 | Unknown | Set of signatures observed in 59 whole-exome sequenced CRCs | Alexandrov *et al.* , 2020 |
| 14 | SBS40 | Unknown | Set of signatures observed in 59 whole-exome sequenced CRCs | Alexandrov *et al.* , 2020 |
| 15 | SBS44 | Defective DNA mismatch repair | Set of signatures observed in 59 whole-exome sequenced CRCs | Alexandrov *et al.* , 2020; Everall *et al.* , 2023 |
| 16 | SBS88 | Colibactin exposure (E. coli bacteria carrying pks pathogenicity island) | Observed in CRCs caused by *Escherichia coli* producing the genotoxin colibactin | Pleguezuelos-Manzano et al., 2020; Everall *et al.* , 2023 |

*Abbreviations: SBS, single base substitution; ID, small insertion/deletion; CRC, colorectal cancer.*

[1] The mutational signatures and proposed aetiologies listed here are part of the SBS and ID mutational signature spectrum as published by Tate *et al.* (COSMIC v3.2).

**Supplementary Table S2.** Participants and their germline *MUTYH* (NM_001128425.1) and *NTHL1* (NM_002528.7) variants identified from clinical diagnostic or research testing included in this study.

| # | Patient ID | Study | AgeDx | Sex | MMR | Tissue tested | Gene | Germline variants | State | ClinVar classification | Genotype |
|---|---|---|---|---|---|---|---|---|---|---|---|
| 1 | Pat_307 | CCFR | 62 | M | pMMR | 3x adenoma, 3x CRC | *MUYTH* | c.536A>G p.(Tyr179Cys) | Homozygous | (Likely) pathogenic | Biallelic *MUTYH* case |
| 2 | Rel_307 | CCFR | 56 | F | pMMR | 1x adenoma, 2x CRC | *MUYTH* | c.536A>G p.(Tyr179Cys) | Homozygous | (Likely) pathogenic | Biallelic *MUTYH* case |
| 3 | Pat_231 | CCFR | 54 | M | pMMR | 1x CRC | *MUYTH* | c.545G>A p.(Arg182His) \| c.536A>G p.(Tyr179Cys) | Compound heterozygous | Pathogenic \| (Likely) pathogenic | Biallelic *MUTYH* case |
| 4 | Pat_357 | CCFR | 64 | M | pMMR | 2x CRC | *MUYTH* | c.1187G>A p.(Gly396Asp) | Homozygous | (Likely) pathogenic | Biallelic *MUTYH* case |
| 5 | Pat_608 | CCFR | 33 | M | pMMR | 2x adenoma, 1x CRC | *MUYTH* | c.1147del p.(Ala385ProfsTer23) | Homozygous | (Likely) pathogenic | Biallelic *MUTYH* case |
| 6 | Pat_301 | CCFR | 50 | F | pMMR | 1x CRC | *MUYTH* | c.536A>G p.(Tyr179Cys) | Homozygous | (Likely) pathogenic | Biallelic *MUTYH* case |
| 7 | Pat_301 | CCFR | 50 | F | dMMR/pMMR | 2x CRCs | *MUYTH* | c.536A>G p.(Tyr179Cys) | Homozygous | (Likely) pathogenic | Biallelic *MUTYH* case |
| 8 | Pat_315 | CCFR | 39 | M | dMMR | 1x CRC | *MUYTH* | c.1187G>A p.(Gly396Asp) | Homozygous | (Likely) pathogenic | Biallelic *MUTYH* case |
| 9 | Pat_822 | CCFR | 39 | M | pMMR | 1x CRC | *MUYTH* | c.536A>G p.(Tyr179Cys) \| c.734G>A p.(Arg245His) | Compound heterozygous | (Likely) pathogenic \| (Likely) pathogenic | Biallelic *MUTYH* case |
| 10 | Pat_206 | CCFR | 59 | M | pMMR | 1x CRC | *MUYTH* | c.1187G>A p.(Gly396Asp) | Homozygous | (Likely) pathogenic | Biallelic *MUTYH* case |
| 11 | Pat_041 | GCPS | 33 | M | pMMR | 1x CRC | *MUYTH* | c.1147del p.(Ala385ProfsTer23) | Homozygous | (Likely) pathogenic | Biallelic *MUTYH* case |
| 12 | Pat_509 | GCPS | 65 | F | pMMR | 2x adenoma | *MUYTH* | c.536A>G p.(Tyr179Cys) | Homozygous | (Likely) pathogenic | Biallelic *MUTYH* case |
| 13 | Pat_706 | GCPS | 73 | F | pMMR | 1x adenoma | *MUYTH* | c.1187G>A p.(Gly396Asp) \| c.933+3A>C p.? | Compound heterozygous | (Likely) pathogenic \| Pathogenic | Biallelic *MUTYH* case |
| 14 | Pat_763 | GCPS | 55 | M | pMMR | 4x adenoma, 1x CRC | *MUYTH* | c.1187G>A p.(Gly396Asp) \| c.533G>C p.(Gly178Ala) | Compound heterozygous | Pathogenic \| Variant of uncertain significance | Suspected biallelic *MUTYH* case |
| 15 | Rel_357 | CCFR | 64 | F | pMMR | 1x CRC | *MUYTH* | c.1187G>A p.(Gly396Asp) | Heterozygous | (Likely) pathogenic | Monoallelic *MUTYH* case |
| 16 | Pat_036 | CCFR | 35 | F | pMMR | 1x CRC | *MUYTH* | c.1187G>A p.(Gly396Asp) | Heterozygous | (Likely) pathogenic | Monoallelic *MUTYH* case |
| 17 | Pat_400 | CCFR | 49 | M | pMMR | 1x CRC | *MUYTH* | c.1187G>A p.(Gly396Asp) | Heterozygous | (Likely) pathogenic | Monoallelic *MUTYH* case |
| 18 | Pat_018 | ANGELS | 39 | F | pMMR | 1x CRC | *MUYTH* | c.1187G>A p.(Gly396Asp) | Heterozygous | (Likely) pathogenic | Monoallelic *MUTYH* case |
| 19 | Pat_427 | GCPS | 57 | M | pMMR | 3x adenoma | *NTHL1* | c.244C>T p.(Gln82Ter) | Homozygous | (Likely) pathogenic | Biallellelic *NTHL1* case |
| 20 | Pat_445 | ANGELS | 53 | F | pMMR | 2x adenoma | *NTHL1* | c.835C>T p.(Gln279Ter) \| c.244C>T p.(Gln82Ter) | Compound heterozygous | (Likely) pathogenic \| (Likely) pathogenic | Biallellelic *NTHL1* case |
| 21 | Rel_445 | ANGELS | 56 | F | pMMR | 2x adenoma | *NTHL1* | c.835C>T p.(Gln279Ter) \| c.244C>T p.(Gln82Ter) | Compound heterozygous | (Likely) pathogenic \| (Likely) pathogenic | Biallellelic *NTHL1* case |
| 22 | Pat_469 | GCPS | 76 | F | pMMR | 1x adenoma, 1x CRC | *NTHL1* | c.244C>T p.(Gln82Ter) | Homozygous | (Likely) pathogenic | Biallellelic *NTHL1* case |
| 23 | Pat_005 | CCFR | 61 | F | pMMR | 1x adenoma, 1x CRC | *NTHL1* | c.244C>T p.(Gln82Ter) \| c.211dup p.(Ala71GlyfsTer2) | Compound heterozygous | (Likely) pathogenic \| (Likely) pathogenic | Biallellelic *NTHL1* case |
| 24 | Pat_110 | CCFR | 61 | F | pMMR | 1x CRC | *NTHL1* | c.835C>T p.(Gln279Ter) \| c.244C>T p.(Gln82Ter) | Heterozygous | (Likely) pathogenic | Monoallellic *NTHL1* case |
| 25 | Pat_108 | CCFR | 43 | M | pMMR | 1x CRC | *NTHL1* | c.244C>T p.(Gln82Ter) | Heterozygous | (Likely) pathogenic | Monoallellic *NTHL1* case |

*Abbreviations: ID, identification number; Pat, patient; Rel, relative; CCFR, Colon Cancer Family Registry; ANGELS, Applying Novel Genomic approaches to Early-onset and suspected Lynch Syndrome colorectal and endometrial cancers; GCPS, The Genetics of Colonic Polyposis Study; CRC, colorectal cancer; MMR, DNA mismatch repair; pMMR, DNA mismatch repair proficient; dMMR, DNA mismatch repair deficient.*
